# Individual-level and neighborhood-level factors associated with longitudinal changes in cardiometabolic measures in participants of a care coordination program

**DOI:** 10.1101/2021.01.25.21250410

**Authors:** Sonal J Patil, Mojgan Golzy, Angela Johnson, Yan Wang, Jerry C Parker, Debra Haire-Joshu, David R Mehr, Randi E Foraker, Robin L Kruse

## Abstract

**Introduction:** Identifying individual and neighborhood-level factors associated with less improvement or worsening cardiometabolic measures in participants of clinic-based interventions may help identify candidates for supplementary community-based interventions.

**Methods:** *Study design:* Secondary data analysis of data from care coordination program cohort, **L**everaging **I**nformation Technology to **G**uide **H**igh **T**ech, **H**igh **T**ouch Care (LIGHT^2^). *Participants:* Medicare, Medicaid, or dual-eligible adults from the University of Missouri Health System enrolled in LIGHT^2^. *Intervention:* Nurse-led care coordination in ten primary care clinics. *Outcomes*: Hemoglobin A1C, low-density-lipoprotein (LDL) cholesterol, and blood pressure. Multivariable generalized linear regression models assessed changes in outcomes after adjusting for clinical and sociodemographic factors, neighborhood-level factors, and driving time to primary care clinics.

**Results:** 6378 participants had pre-and post-intervention cardiometabolic measures reported (61% women, 86.3% White, non-Hispanic ethnicity, mean age 62.7 [SD, 18.5] years). In adjusted models, pre-intervention measures and female gender were associated with worsening of all cardiometabolic measures. Women’s LDL-cholesterol worsened compared to men irrespective of pre-intervention levels (β 7.87, 95% CI 5.24 to 10.5, p<.001). Women with hemoglobin A1C> 6.8% had worsening hemoglobin A1C compared to men (main effect β −1.28, 95% CI −1.95 to −0.61, p<.001; interaction effect β 0.19, 95% CI 0.09 to 0.28, p<.001). Women with systolic blood pressure >121 mm of Hg had worsening diastolic blood pressure compared to men (main effect β −5.42, 95% CI −9.8 to 0.098, p = 0.016; interaction effect β 0.04, 95% CI 0.01 to 0.078, p = 0.009). Adding neighborhood-level factors or driving time to primary care clinics did not improve the overall fit of the models.

**Conclusions:** In a solely clinic-based care coordination program, increasing baseline cardiometabolic measures and female gender were associated with worsening cardiometabolic outcomes. Further research to understand the causes of these associations may help tailor clinic-community-linked interventions.

## Introduction

Cardiovascular disease (CVD) is the leading cause of death in the US, and cardiometabolic risk factors for CVD are prevalent and well-known.[1, 2] Despite the availability of effective treatments, significant disparities in CVD and cardiometabolic outcomes persist.[3-6] Care coordination can deliver better access and quality of care, and community-based interventions can empower patients to address social and behavioral determinants of cardiometabolic outcomes.[7-12] However, care coordination research has primarily focused on healthcare costs with inconsistent results.[13-18] On the other hand, clinical translation of successful multilevel clinic-community-linked interventions that reduce cardiometabolic health disparities has been challenging.[9, 19-21] Most primary care practices do not have time or resources to offer widespread care coordination, individual-level social risk screenings, and community-based care.[20, 22-26] Identifying those who do not benefit from clinic-based interventions can help focus clinic and community-based resources.

We sought to identify potential candidates and high burden communities with cardiometabolic disparities for community-based interventions.[11] We aimed to identify individual and neighborhood-level factors associated with less improvement or worsening cardiometabolic outcomes after participation in a 2-year clinic-based nurse-led care coordination program. We hypothesized that worsening cardiometabolic measures would be associated with living in an area with higher social risks or longer driving time to primary care clinic locations.

## Methods

We performed a secondary analysis of data from a prospective cohort of University of Missouri Healthcare (MUHC) system patients enrolled in the **L**everaging **I**nformation Technology to **G**uide **H**igh **T**ech, **H**igh **T**ouch Care (LIGHT^2^) project from July 1, 2013, to June 30, 2015.[27] The University of Missouri institutional review board determined the LIGHT^2^ program to be a quality improvement activity not requiring institutional review board review. We used the Strengthening the Reporting of Observational Studies in Epidemiology (STROBE) reporting guidelines for this study.[28]

### Study design and setting

Funded with a Centers for Medicare and Medicaid Services (CMS) innovation award, the LIGHT^2^ program was a combination of information technology components (High Tech) and health care coordination by nurse care managers (High Touch).[27, 29] The High Tech component included dashboards and a patient portal for communication with physicians and nurse care managers. The High Touch component was care coordination provided by nurse care managers; 25 nurse care managers worked in 10 family and community medicine and internal medicine clinics providing services. Patients were assigned tiers based on their chronic disease diagnoses and health care chronic diseases or 1 stable chronic condition, respectively. Tier 3 and 4 patients had at least 1 chronic condition, 1 or more hospital visits, and several outpatient visits. Nurse care managers provided as-needed education and support for Tier 1 and 2 patients regarding new chronic disease diagnoses or worsening of chronic conditions; Tier 3 and 4 patients were contacted monthly. Hence, more patients in Tiers 3 and 4 (80%) received nurse coordination services compared to patients in Tiers 1 and 2 (64%). A documentation system measuring the frequency and time of nurse care manager contacts was created using the Agency of Healthcare Research and Quality (AHRQ) care coordination atlas.[29, 31] The study compiled data on claims and participants’ electronic health record (EHR) data, including healthcare utilization, clinic visits, basic demographic data, diagnosis, labs, and nurse care coordination contacts, along with geocoded patient addresses. Cohort recruitment started in February 2013; 9932 participants were recruited by July 2013. Deaths and relocations decreased the number of participants to 8593 by March 2015. The goals of the LIGHT^2^ intervention were to achieve net cost savings, increase preventive service use, and improve the management of chronic diseases. The final evaluation of the program showed higher spending than the comparison group, and most patients received recommended preventive services.[27] The cardiometabolic measures monitored during LIGHT^2^ intervention were hemoglobin A1C (HbA1C), blood pressure (BP), and low density-lipoprotein (LDL) cholesterol. Primary results showed minimal changes in cardiometabolic outcomes. Detailed results of primary outcomes, nurse care coordination implementation, and fidelity of LIGHT^2^ have been previously published.[27, 29]

### Participants

Participants were all Medicare, Medicaid, or dual-eligible patients receiving primary care services in any of 10 family and community medicine or internal medicine clinics in the MUHC system during the project period. Because we examined factors associated with a change in cardiometabolic measures in the presence of nurse-led care coordination, we included only participants with HbA1C, LDL cholesterol, or BP reported before and after the intervention completion.

### Outcomes

We extracted HbA1C, BP, and LDL-cholesterol levels before and after the LIGHT^2^ program. Recruitment was complete and the care coordination documentation system was implemented on 1 July, 2013. We used measures reported within 6 months before that date or before the first nurse care manager contact within the first 3 months of the intervention as the pre-intervention values. For post-intervention measures, we used the values reported within 6 months after the intervention completion date. If there were no values reported in the 6 months after the completion of the intervention, we used the last reported value within the last 3 months of the intervention. We used the average of the last 2 reported BP readings if there were multiple readings in the defined pre-intervention and post-intervention time-period. Individuals with very low pre-intervention values were not included in the analyses. More specifically, if pre-intervention LDL<60 mg/dL, pre-intervention HbA1C<5.5%, pre-intervention diastolic BP (DBP)<40 mm of Hg, and pre-intervention systolic BP (SBP)<90 mm of Hg, individuals were excluded. We also excluded 2 extremely high values (above 350) of post-intervention LDL cholesterol. The population comprised of 6378 individuals after exclusions.

### Variables

We included clinical and geospatial variables accounting for social and behavioral domains and measures recommended by the National Academy of Medicine (NAM).[32] We identified clinical and area-level variables easily extracted from routinely collected clinical and sociodemographic data in the EHR. The NAM recommended domains include race and ethnicity, which were extracted from EHR; education, which we extracted at neighborhood-level; financial resource strain was extracted as poverty at neighborhood-level and access to healthy food as the distance to nearest grocery store; stress and depression were denoted by the presence of mood disorders from EHR; physical activity opportunities were extracted at neighborhood-level; tobacco and alcohol status extracted from EHR; social connection or isolation measured by marital status in EHR and neighborhood-level social capital; intimate partner violence measured by neighborhood-level domestic violence injury rates. As the LIGHT^2^ program was focused on nurse care coordination for healthcare utilization and chronic disease management, we adjusted for health resource utilization, nurse care manager contacts, and the number of comorbidities. Additionally, as LIGHT^2^ program was solely based in primary care clinics, we examined driving time to the primary care clinic from the patient’s geocoded residential address. As baseline cardiometabolic health, including BMI, are used by clinicians for CVD risk stratification, we adjusted for baseline values of cardiometabolic measures.

We included basic demographic data such as age, sex, race/ethnicity, as well as smoking status, alcohol status, and geocoded residential addresses from the EHR. We extracted the total number of comorbidities, nurse care manager contacts, and presence or absence of a mood disorder reported as of the last day of intervention based on the International Classification of Diseases, Ninth Revision, Clinical Modification diagnosis codes (ICD 9 codes). We measured healthcare utilization and defined high-resource utilizers as individuals with 4 or more emergency room visits, or 2 or more inpatient encounters in the previous 12 months. The patient’s residential address and primary care physician’s (PCP) clinic addresses were geocoded using ArcGIS Online World Geocoding Service.[33] If a patient was seen at 2 or more primary care clinic locations during the intervention period, we coded the distance to the clinic as missing to avoid misattributions. Distance variables were calculated using an origin-destination cost matrix network analysis using a StreetMap North America street network dataset.[34] Supermarket locations were obtained from the Reference USA US Businesses dataset.[35, 36] Socioeconomic status of patients’ neighborhoods was extracted from the US Census Bureau 2010-2014 American Community Survey (ACS) 5-Year Estimates.[37] The percentage of population <200% federal poverty level (FPL) was extracted at patient’s census-tract level, and the percentage of population without high school diploma was determined at census block group-level.[38] Neighborhood (ZIP-code level) social capital was assessed using the number of civic or social organizations per capita, obtained by summarizing data from the 2017 US Census Bureau ZIP code Business Patterns.[39] Domestic violence injury rates were extracted at Zip-code level. To assess opportunities for physical activity, we generated WalkScore™ values for each patient’s census block.[40] Visual assessment found that the WalkScore™ dataset may not represent physical activity opportunities in areas with networks of unpaved trails such as our local county and rural areas; hence, we excluded these measures. See S1 File for detailed information of included neighborhood level variables.

## Statistical methods

After adjusting for covariates, we examined the changes in HbA1C, LDL, and BP from before the 1 July, 2013 nurse-led care coordination start date to after the end of the project in June 2015. Dataset was created with clinical, sociodemographic, and geospatial variables followed by data analysis between November 2019 and October 2020. We fitted a separate generalized linear model (GLM) for each of these measures as the dependent variable. We adjusted for covariates, including pre-intervention risk factor values and pre-intervention body mass index (BMI). The GLM model assumptions were checked. Using the Pearson correlation between continuous variables, we chose one variable from each group of variables. Variance Inflation Factors (VIF) were calculated as 1/ (type I tolerance) of the GLM model for categorical variables. The cutoff point was 5. The overall fit of the models was assessed using the coefficient of determinations (R^2^). We report statistical significance at p < .01. Results were generated using SAS software.

### Model 1

Included covariates readily available in the clinical chart: age, gender, race/ethnicity, marital status, smoking status, alcohol use, healthcare utilization 12 months before the study’s start date, pre-intervention cardiometabolic risk factor levels, pre-intervention BMI, presence of mood disorder, total number of chronic health conditions, and total number of nurse care manager contacts during the study period. Significant interactions between the covariates and the pre-intervention values were included in the model.

### Model 2

Included the covariates in Model 1 and also included neighborhood-level measures of poverty, education, social capital, and domestic violence injury rates.

### Model 3

Included the covariates in Model 2 and added physical determinants of access to healthy food (grocery store) and healthcare (PCP clinic location) from the patient’s geocoded address.

At the census block group level, we noted collinearity among the percentage of population below 200% of the FPL and the Area Deprivation Index.[41] Thus, we included only the percentage of the population below 200% of the FPL in our models. We dichotomized driving time to PCP clinic location to 30 minutes or less, and over 30 minutes, as Health Professional Shortage Area (HPSA) designation utilizes driving time of more than 30 to 40 minutes to the nearest source of care as one of the scoring criteria.[42] Based on frequency distributions and outliers, we represented percentage of the population without a high school diploma and the number of nurse care manager contacts as categorical variables by quartiles.

### Sensitivity analysis

We performed 2 additional analyses for each outcome: 1) We used a backward-selection technique to remove non-significant factors step-by-step to obtain a parsimonious model with only significant factors, and 2) we fit a mixed-effects model to test for random variability across clinics.

## Results

Of 8593 eligible participants in the LIGHT^2^ cohort, 6378 participants had at least 1 cardiometabolic measure reported both before and after the intervention. Figure 1 Flow Diagram illustrates the derivation of participants for each measure. The cohort description is included in Table 1. Because our cohort was primarily White, non-Hispanic ethnicity (86.3%), we dichotomized the race/ethnicity variable.

**Table 1.**
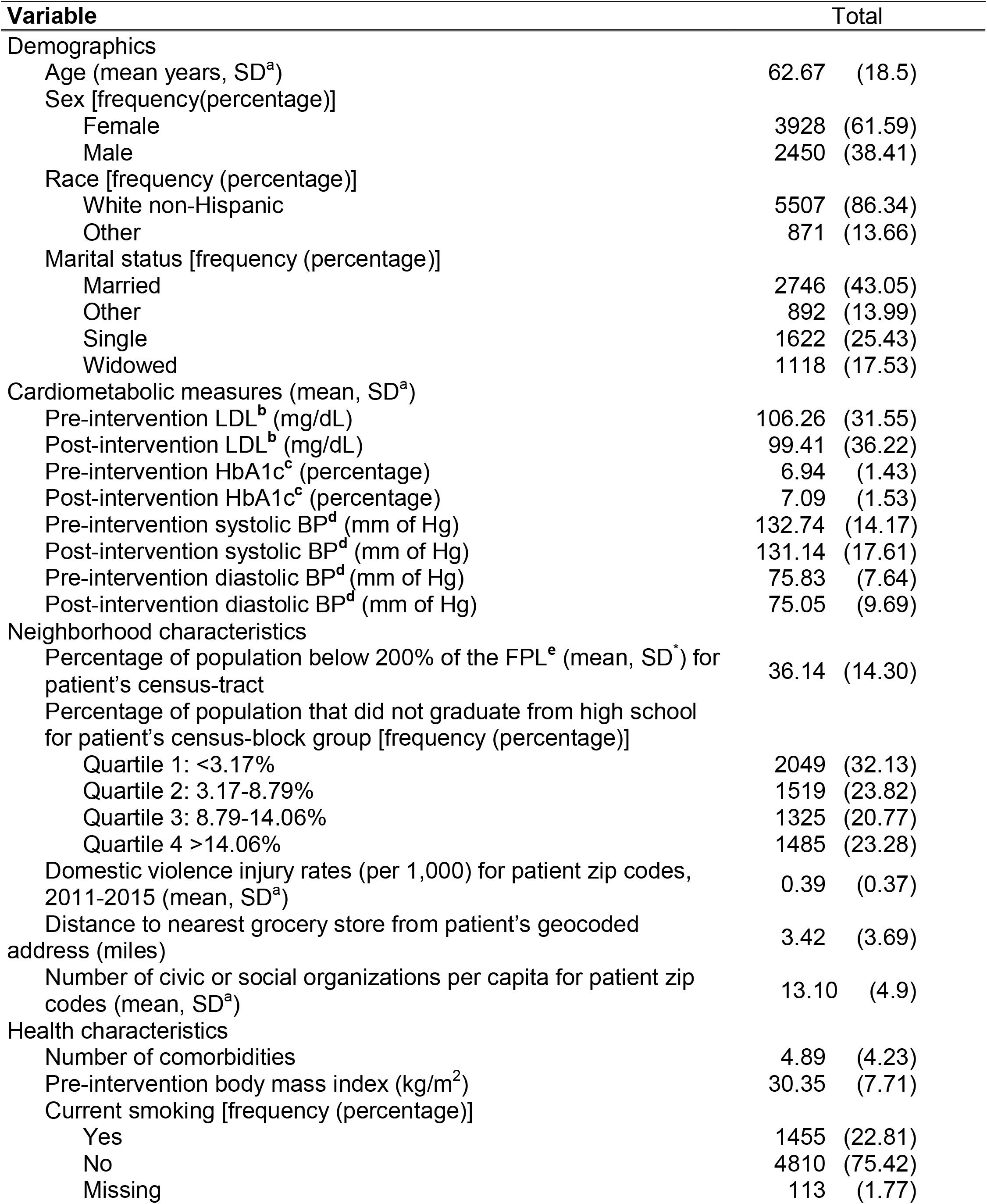
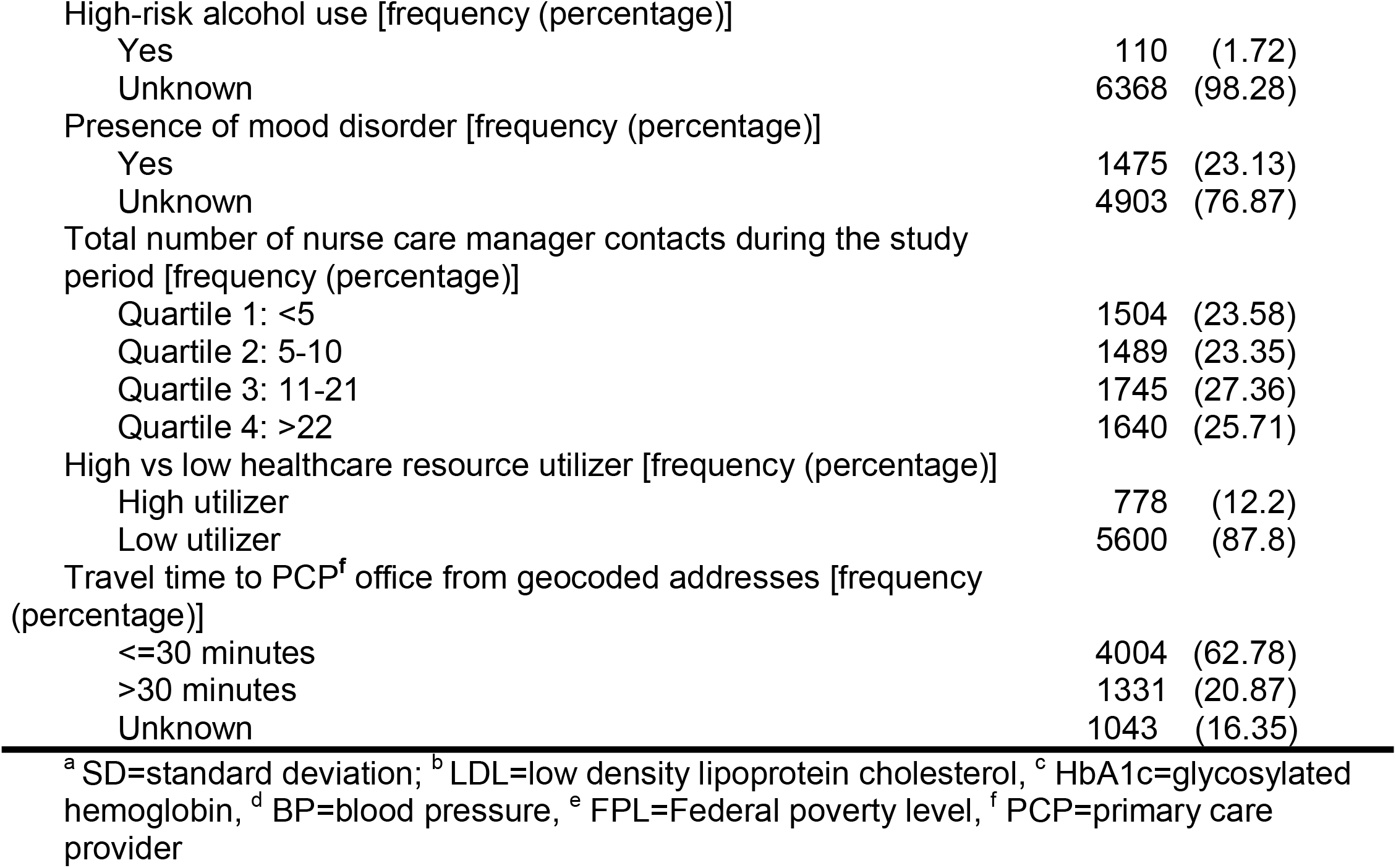
Characteristics of the study sample (N=6378)

**Figure 1.**
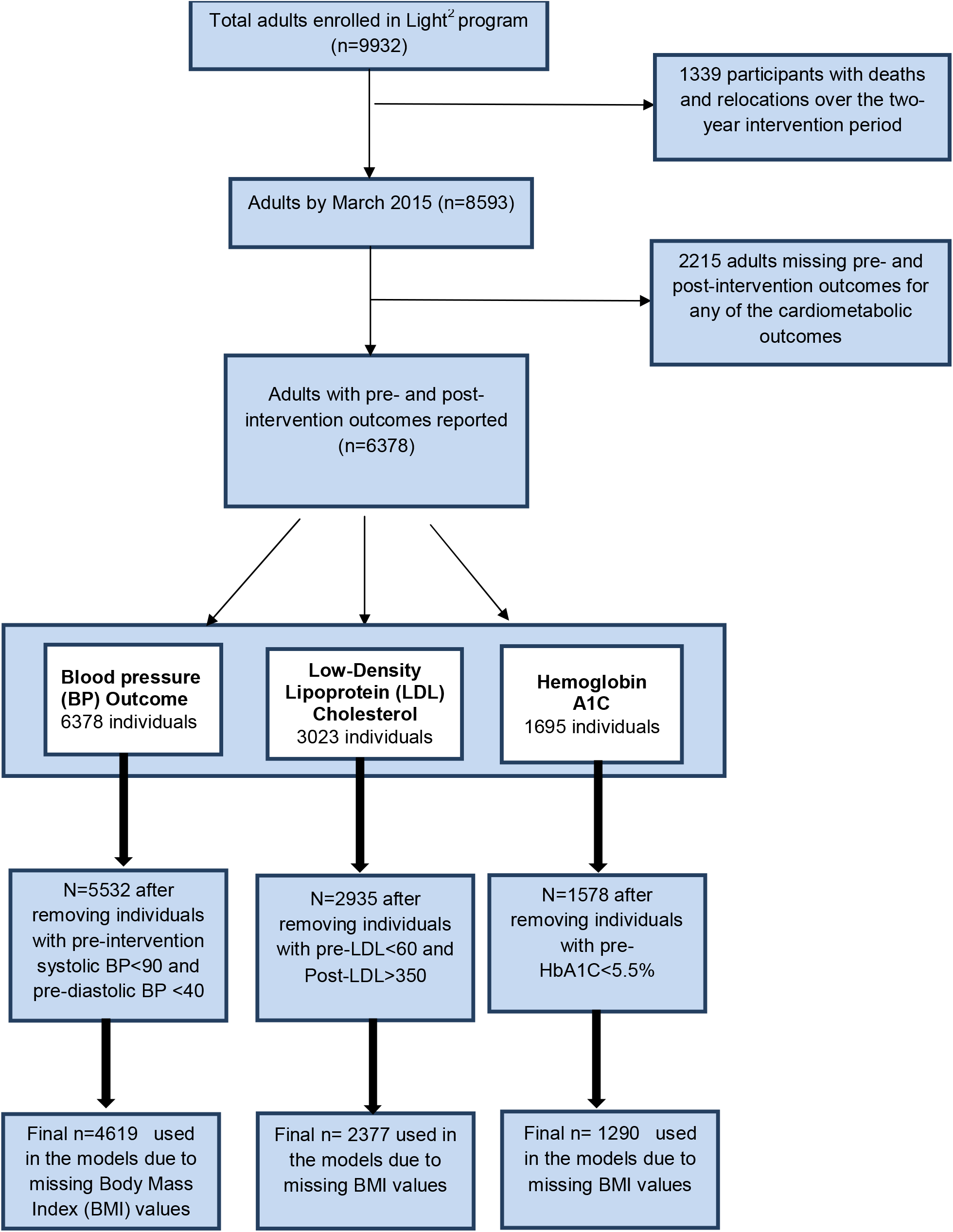
Flow diagram of participants Flow diagram of <u>**L**</u>everaging <u>**I**</u>nformation Technology to <u>**G**</u>uide <u>**H**</u>igh <u>**T**</u>ech, <u>**H**</u>igh <u>**T**</u>ouch Care (LIGHT^2^) program participants and final number of participants included in analysis for each cardiometabolic measure

We had LDL cholesterol outcomes for 2377 participants. See Table 2 for detailed results and R^2^ for all models. The final multivariable model for LDL cholesterol showed significant deterioration associated with pre-intervention LDL cholesterol (β 0.56, 95% CI 0.52 to 0.597, p<.001) and female gender compared to males (β 7.87, 95% CI 5.24 to 10.5, p<.001). Significant improvement in LDL cholesterol was associated with increasing age (β −0.28, 95% CI −0.4 to −0.16, p<.001) and higher area-level domestic violence injury rates (β −5.41, 95% CI −9.08 to −1.75, p = 0.004)

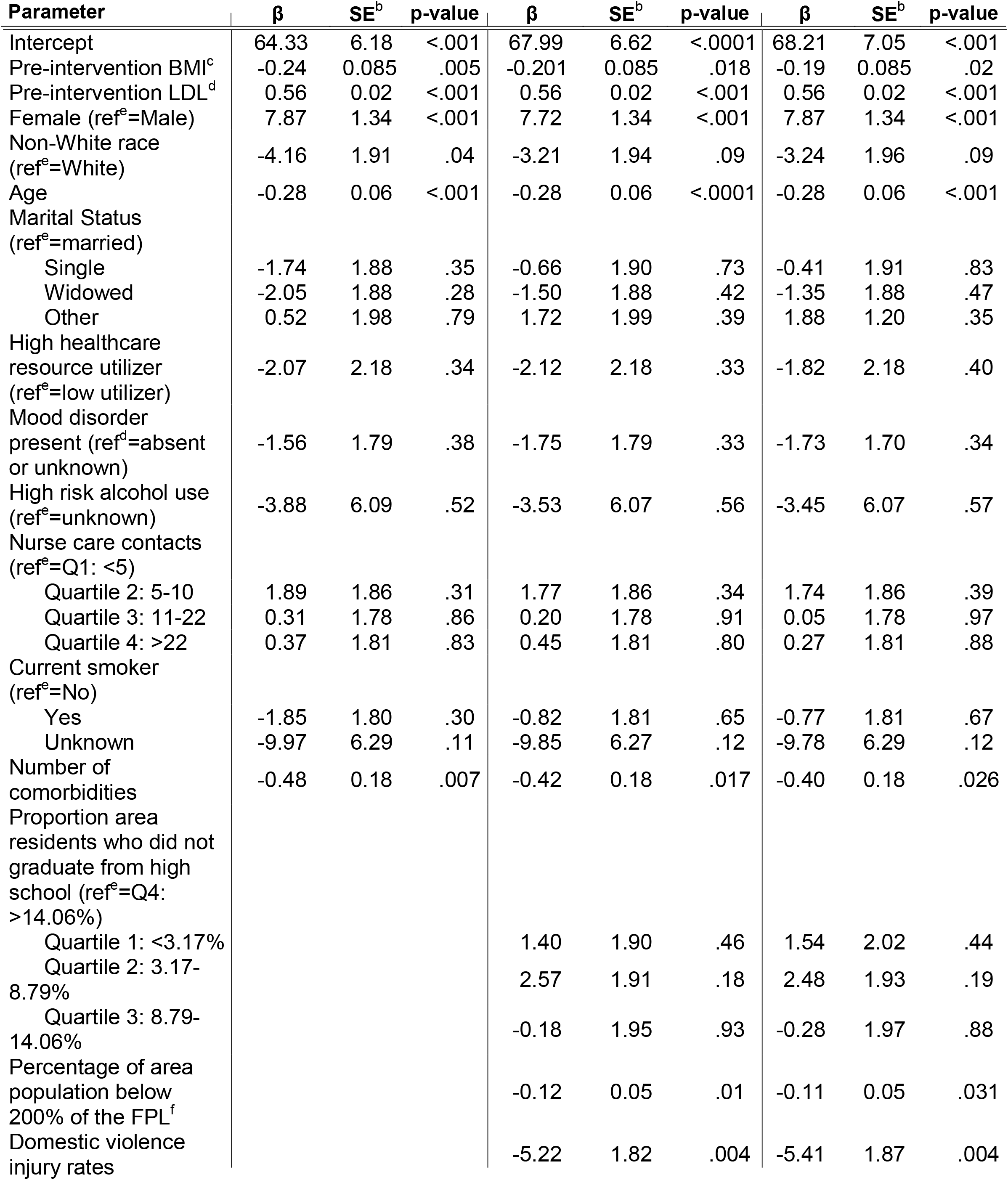
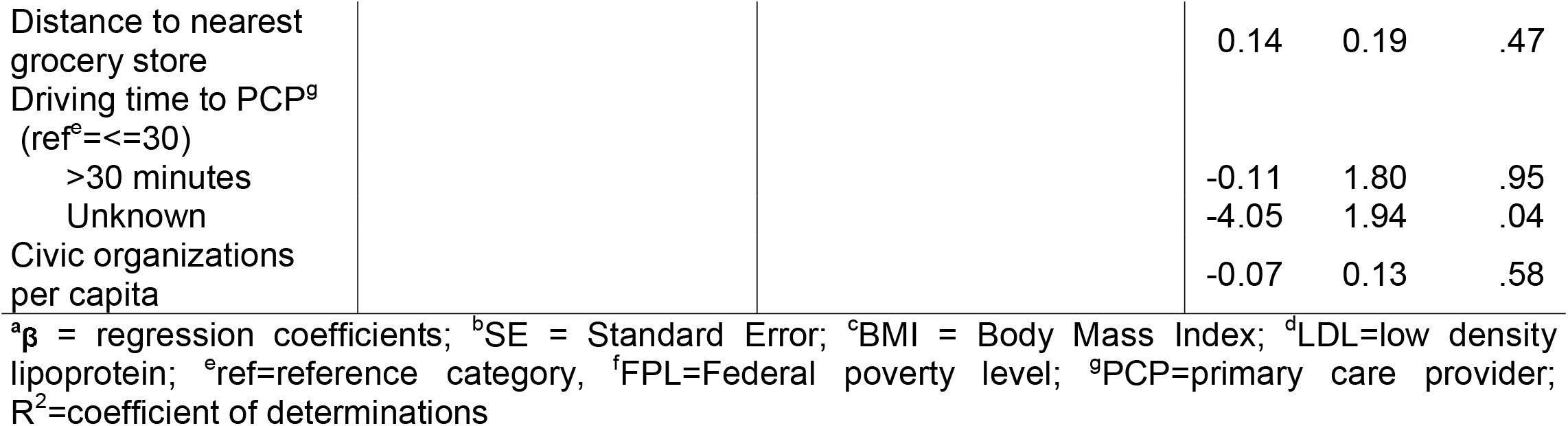

|  |  |  |  |  |  |  |  |  |  |
| --- | --- | --- | --- | --- | --- | --- | --- | --- | --- |
| Intercept | 64.33 | 6.18 | <.001 | 67.99 | 6.62 | <.0001 | 68.21 | 7.05 | <.001 |
| Pre-intervention BMI<sup>c</sup> | -0.24 | 0.085 | .005 | -0.201 | 0.085 | .018 | -0.19 | 0.085 | .02 |
| Pre-intervention LDL<sup>d</sup> | 0.56 | 0.02 | <.001 | 0.56 | 0.02 | <.001 | 0.56 | 0.02 | <.001 |
| Female (ref<sup>e</sup>=Male) | 7.87 | 1.34 | <.001 | 7.72 | 1.34 | <.001 | 7.87 | 1.34 | <.001 |
| Non-White race<br>(ref<sup>e</sup>=White) | -4.16 | 1.91 | .04 | -3.21 | 1.94 | .09 | -3.24 | 1.96 | .09 |
| Age | -0.28 | 0.06 | <.001 | -0.28 | 0.06 | <.0001 | -0.28 | 0.06 | <.001 |
| Marital Status<br>(ref<sup>e</sup>=married) |  |  |  |  |  |  |  |  |  |
| Single | -1.74 | 1.88 | .35 | -0.66 | 1.90 | .73 | -0.41 | 1.91 | .83 |
| Widowed | -2.05 | 1.88 | .28 | -1.50 | 1.88 | .42 | -1.35 | 1.88 | .47 |
| Other | 0.52 | 1.98 | .79 | 1.72 | 1.99 | .39 | 1.88 | 1.20 | .35 |
| High healthcare<br>resource utilizer<br>(ref<sup>e</sup>=low utilizer) | -2.07 | 2.18 | .34 | -2.12 | 2.18 | .33 | -1.82 | 2.18 | .40 |
| Mood disorder<br>present (ref<sup>d</sup>=absent<br>or unknown) | -1.56 | 1.79 | .38 | -1.75 | 1.79 | .33 | -1.73 | 1.70 | .34 |
| High risk alcohol use<br>(ref<sup>e</sup>=unknown) | -3.88 | 6.09 | .52 | -3.53 | 6.07 | .56 | -3.45 | 6.07 | .57 |
| Nurse care contacts<br>(ref<sup>e</sup>=Q1: <5) |  |  |  |  |  |  |  |  |  |
| Quartile 2: 5-10 | 1.89 | 1.86 | .31 | 1.77 | 1.86 | .34 | 1.74 | 1.86 | .39 |
| Quartile 3: 11-22 | 0.31 | 1.78 | .86 | 0.20 | 1.78 | .91 | 0.05 | 1.78 | .97 |
| Quartile 4: >22 | 0.37 | 1.81 | .83 | 0.45 | 1.81 | .80 | 0.27 | 1.81 | .88 |
| Current smoker<br>(ref<sup>e</sup>=No) |  |  |  |  |  |  |  |  |  |
| Yes | -1.85 | 1.80 | .30 | -0.82 | 1.81 | .65 | -0.77 | 1.81 | .67 |
| Unknown | -9.97 | 6.29 | .11 | -9.85 | 6.27 | .12 | -9.78 | 6.29 | .12 |
| Number of<br>comorbidities | -0.48 | 0.18 | .007 | -0.42 | 0.18 | .017 | -0.40 | 0.18 | .026 |
| Proportion area<br>residents who did not<br>graduate from high<br>school (ref<sup>e</sup>=Q4:<br>>14.06%) |  |  |  |  |  |  |  |  |  |
| Quartile 1: <3.17% |  |  |  | 1.40 | 1.90 | .46 | 1.54 | 2.02 | .44 |
| Quartile 2: 3.17-8.79% |  |  |  | 2.57 | 1.91 | .18 | 2.48 | 1.93 | .19 |
| Quartile 3: 8.79-14.06% |  |  |  | -0.18 | 1.95 | .93 | -0.28 | 1.97 | .88 |
| Percentage of area<br>population below<br>200% of the FPL<sup>f</sup> |  |  |  | -0.12 | 0.05 | .01 | -0.11 | 0.05 | .031 |
| Domestic violence<br>injury rates |  |  |  | -5.22 | 1.82 | .004 | -5.41 | 1.87 | .004 |

|  |  |  |  |  |  |
| --- | --- | --- | --- | --- | --- |
| Distance to nearest grocery store |  |  | 0.14 | 0.19 | .47 |
| Driving time to PCP<sup>g</sup> (ref<sup>e</sup>=<=30) |  |  |  |  |  |
| >30 minutes |  |  | -0.11 | 1.80 | .95 |
| Unknown |  |  | -4.05 | 1.94 | .04 |
| Civic organizations per capita |  |  | -0.07 | 0.13 | .58 |

<sup>a</sup> $\beta$ = regression coefficients; <sup>b</sup>SE = Standard Error; <sup>c</sup>BMI = Body Mass Index; <sup>d</sup>LDL=low density lipoprotein; <sup>e</sup>ref=reference category, <sup>f</sup>FPL=Federal poverty level; <sup>g</sup>PCP=primary care provider; R<sup>2</sup>=coefficient of determinations

The multivariable models for SBP (S1 Table) showed significant worsening with higher BMI (β 0.09, 95% CI 0.043 to 0.147, p<.001), higher pre-intervention SBP (β 0.93, 95% CI 0.79 to 1.06, p<.001), and higher area-level interpersonal injury rates (β 2.04, 95% CI 0.93 to 3.16, p<.001). The interaction between female gender and pre-intervention SBP levels (main effect β −8.93, 95% CI −16.7 to −1.19 p =0.02; interaction effect β 0.067, 95% CI 0.008 to 0.124, p = 0.02) indicates worsening trend in SBP for women compared to men with pre-intervention SBP >134 mm Hg but results do not reach the set statistical significance (S1 Figure). The final model for DBP (Table 3) showed worsening of DBP associated with increasing BMI (β 0.078, 95% CI 0.048 to 0.108, p<.001), pre-intervention SBP (β 0.15, 95% CI 0.078 to 0.23, p<.001), pre-intervention DBP (β 0.822, 95% CI 0.749 to 0.909, p<.001), smoking versus non-smoking (β 0.80, 95% CI 0.22 to 1.38, p = 0.007), known hazardous alcohol use (β 2.44, 95% CI 0.76 to 4.11, p = 0.004), and area-level domestic violence injury rates (β 1.01, 95% CI 0.37 to 1.65, p = 0.002). There also was worsening DBP noted for women compared to men with pre-intervention SBP >121 mm Hg (main effect β −5.42, 95% CI - 9.8 to 0.098, p = 0.016; interaction effect β 0.04, 95% CI 0.01 to 0.078, p = 0.009) (S2 Figure). Age was associated with decreasing SBP when pre-intervention SBP was <153 mm Hg and decreasing DBP when pre-intervention SBP was < 119 mm Hg. A significant interaction of the percentage of population <200% FPL with pre-intervention DBP was observed in the SBP and DBP outcomes models. The association of the increasing percentage of population <200% FPL switches from worsening to small improvements in SBP and DBP at pre-intervention DBP cutoff point of 75mm Hg and 78 mm Hg, respectively.

**Table 3:** Multivariable models of diastolic blood pressure for 4619 participants with pre-and post-intervention measures.

|  | Model 1 (R<sup>2</sup> 0.39) |  |  | Model 2 (R<sup>2</sup> 0.39) |  |  | Model 3 (R<sup>2</sup> 0.39) |  |  |
| --- | --- | --- | --- | --- | --- | --- | --- | --- | --- |
| Parameter | $\beta^a$ | SE<sup>b</sup> | p-value | $\beta^a$ | SE<sup>b</sup> | p-value | $\beta^a$ | SE<sup>b</sup> | p-value |
| Intercept | 0.58 | 4.48 | 0.89 | -7.47 | 5.51 | 0.17 | -7.81 | 5.54 | 0.16 |
| Pre-intervention BMI<sup>c</sup> | 0.08 | 0.015 | <.001 | 0.07 | 0.015 | <.001 | 0.078 | 0.015 | <.001 |
| Pre-intervention SBP<sup>d</sup> | 0.14 | 0.038 | <.001 | 0.15 | 0.038 | 0.001 | 0.15 | 0.039 | <.001 |
| Pre-intervention DBP<sup>e</sup> | 0.73 | 0.019 | <.001 | 0.82 | 0.041 | <.001 | 0.82 | 0.041 | <.001 |
| Female (ref<sup>g</sup>=Male) | -4.89 | 2.26 | 0.03 | -5.43 | 2.26 | 0.016 | -5.42 | 2.26 | 0.016 |
| Pre-intervention SBP<sup>d</sup>*<br>Female | 0.041 | 0.017 | 0.017 | 0.045 | 0.017 | 0.008 | 0.045 | 0.02 | 0.009 |
| Race Other than White<br>(ref<sup>f</sup>=White) | -0.61 | 0.34 | 0.06 | -0.58 | 0.34 | 0.09 | -0.55 | 0.34 | 0.113 |
| Age | 0.28 | 0.06 | <.001 | 0.31 | 0.06 | <.001 | 0.31 | 0.06 | <.001 |
| Pre-intervention SBP<sup>d</sup> *age | -0.002 | 0.0005 | <.001 | -0.002 | 0.0005 | <.001 | -0.002 | 0.0005 | <.001 |
| High Healthcare Resource<br>Utilizer (ref<sup>f</sup> Low Healthcare<br>Resource Utilizer) | -0.10 | 0.33 | 0.76 | -0.054 | 0.33 | 0.89 | -0.063 | 0.33 | 0.85 |
| Q1: <5 Nurse care contacts<br>(ref<sup>f</sup>) |  |  |  |  |  |  |  |  |  |
| Q2: 5-10 Nurse care<br>contacts | -0.60 | 0.33 | 0.07 | -0.55 | 0.32 | 0.09 | -0.56 | 0.32 | 0.08 |
| Q3: 11-21 Nurse<br>care contacts | -0.09 | 0.31 | 0.78 | -0.09 | 0.31 | 0.77 | -0.10 | 0.31 | 0.74 |
| Q4: >22 Nurse care<br>contacts | -0.89 | 0.32 | 0.005 | -0.88 | 0.32 | 0.005 | -0.88 | 0.32 | 0.006 |
| Smoker Yes<br>(ref<sup>f</sup>=No) | 1.01 | 0.29 | <.001 | 0.82 | 0.30 | 0.006 | 0.80 | 0.30 | 0.007 |
| Smoking status missing (ref<sup>f</sup>=No) | -1.20 | 1.02 | 0.24 | -1.25 | 1.01 | 0.22 | -1.23 | 1.01 | 0.22 |
| Marital Status (Married) ref<sup>f</sup> |  |  |  |  |  |  |  |  |  |
| Marital Status Single | 0.016 | 0.33 | 0.96 | -0.005 | 0.33 | 0.99 | 0.018 | 0.33 | 0.95 |
| Marital Status Widowed | -0.35 | 0.33 | 0.28 | -0.40 | 0.33 | 0.22 | -0.39 | 0.33 | 0.23 |
| Marital Status Other | 0.76 | 0.35 | 0.031 | 0.66 | 0.35 | 0.06 | 0.68 | 0.36 | 0.06 |
| Mood Disorder present (ref<sup>f</sup>=Mood Disorder absent or unknown) | -0.37 | 0.29 | 0.19 | -0.39 | 0.29 | 0.18 | -0.39 | 0.29 | 0.18 |
| High risk Alcohol use (ref<sup>f</sup> Alcohol use unknown) | 2.41 | 0.86 | 0.005 | 2.39 | 0.85 | 0.005 | 2.44 | 0.86 | 0.004 |
| Number of comorbidities | 0.08 | 0.03 | 0.005 | 0.07 | 0.03 | 0.010 | 0.07 | 0.03 | 0.011 |
| Proportion not graduated high school in the area, Q4 >14.06% (ref<sup>f</sup>) |  |  |  |  |  |  |  |  |  |
| Q1 <3.17% |  |  |  | -0.68 | 0.33 | 0.041 | -0.59 | 0.35 | 0.08 |
| Q2 3.17-8.79 % |  |  |  | -0.79 | 0.33 | 0.017 | -0.74 | 0.33 | 0.028 |
| Q3 8.79- 14.06% |  |  |  | 0.001 | 0.33 | 0.99 | 0.03 | 0.34 | 0.93 |
| Percentage of area population below 200% of the FPL<sup>g</sup> |  |  |  | 0.18 | 0.076 | 0.014 | 0.19 | 0.07 | 0.014 |
| Pre-intervention DBP<sup>e</sup>Poverty200FPL<sup>g</sup> |  |  |  | -0.002 | 0.001 | 0.015 | -0.0024 | 0.001 | 0.016 |
| Domestic violence injury rates |  |  |  | 0.94 | 0.32 | 0.003 | 1.011 | 0.33 | 0.002 |
| Distance to nearest grocery store |  |  |  |  |  |  | 0.05 | 0.034 | 0.14 |
| Time to PCP<=30 (ref<sup>f</sup>) |  |  |  |  |  |  |  |  |  |
| Time to PCP<sup>h</sup>>30 |  |  |  |  |  |  | -0.26 | 0.31 | 0.40 |
| Time to PCP unknown |  |  |  |  |  |  | 0.16 | 0.31 | 0.60 |
| Civic organizations per capita |  |  |  |  |  |  | 0.012 | 0.02 | 0.61 |

<sup>a</sup>β = Regression coefficient; <sup>b</sup>SE = Standard Error; <sup>c</sup> BMI=body mass index (kg/m<sup>2</sup>), <sup>d</sup> SBP=systolic blood pressure, <sup>e</sup> DBP=diastolic blood pressure, <sup>f</sup> ref=reference category, <sup>g</sup>FPL=Federal poverty level; <sup>h</sup>PCP = primary care provider, <sup>i</sup>R<sup>2</sup>=coefficient of determinations

The multivariable models for HbA1C (Table 4) showed significant deterioration associated with increasing baseline HbA1C (β 0.51, 95% CI 0.43 to 0.586, p<.001). There was a significant interaction between pre-intervention HbA1C and female gender (main effect β −1.28, 95% CI −1.95 to −0.61, p<.001; interaction effect β 0.19, 95% CI 0.09 to 0.28, p<.001), with HbA1C worsening if the pre-intervention HbA1C is more than 6.8% in women compared to men (S3 Figure). Similarly, non-White race/ethnicity was associated with improved HbA1C (main effect β −1.159, 95% CI −1.95 to −0.367, p = .004, interaction effect β 0.14, 95% CI 0.033 to 0.25, p = .011), but the HbA1C worsens for non-White patients with a pre-intervention HbA1C greater than 8.1%. Age and smoking status were associated with improved HbA1C across the intervention.

**Table 4:**
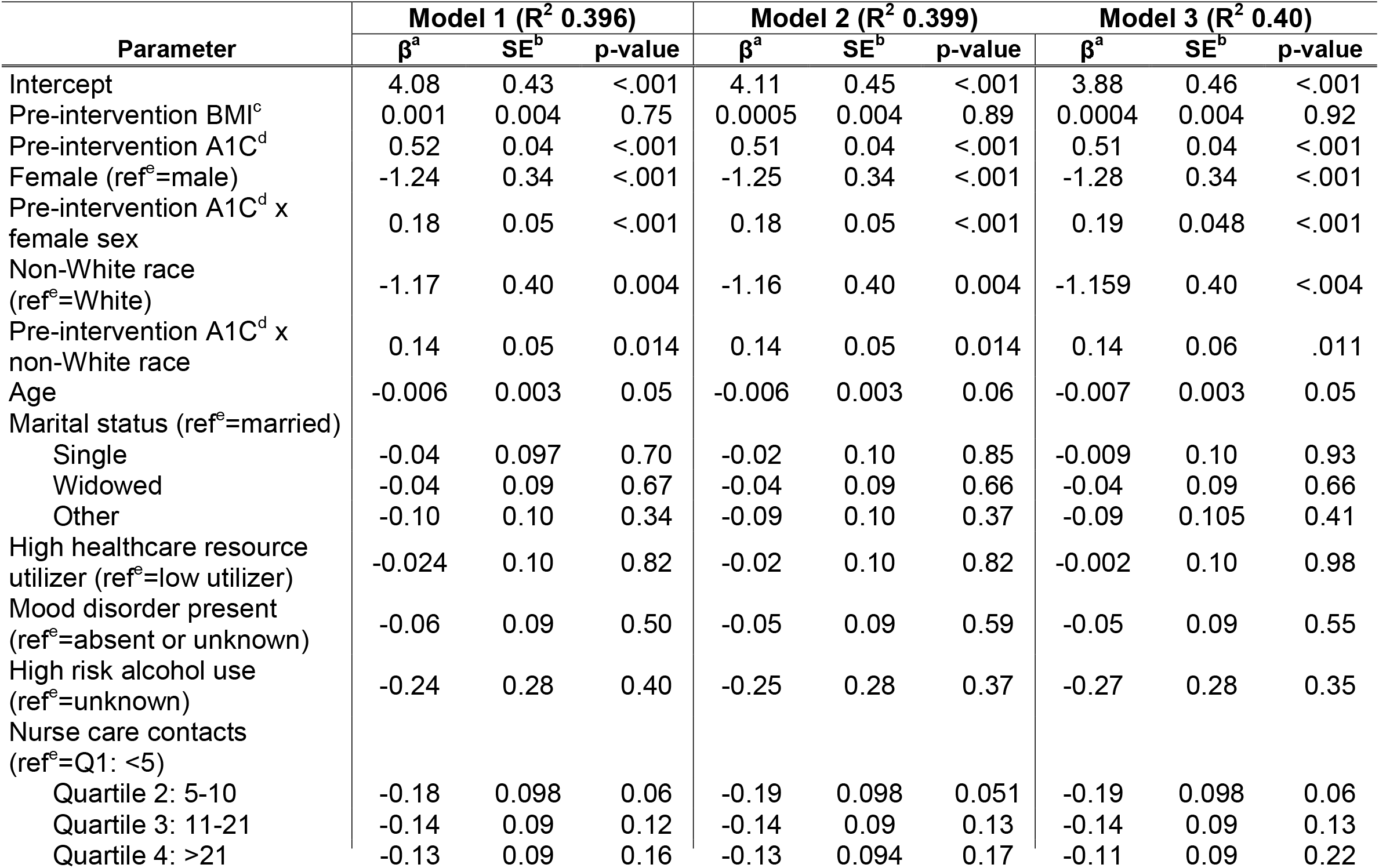
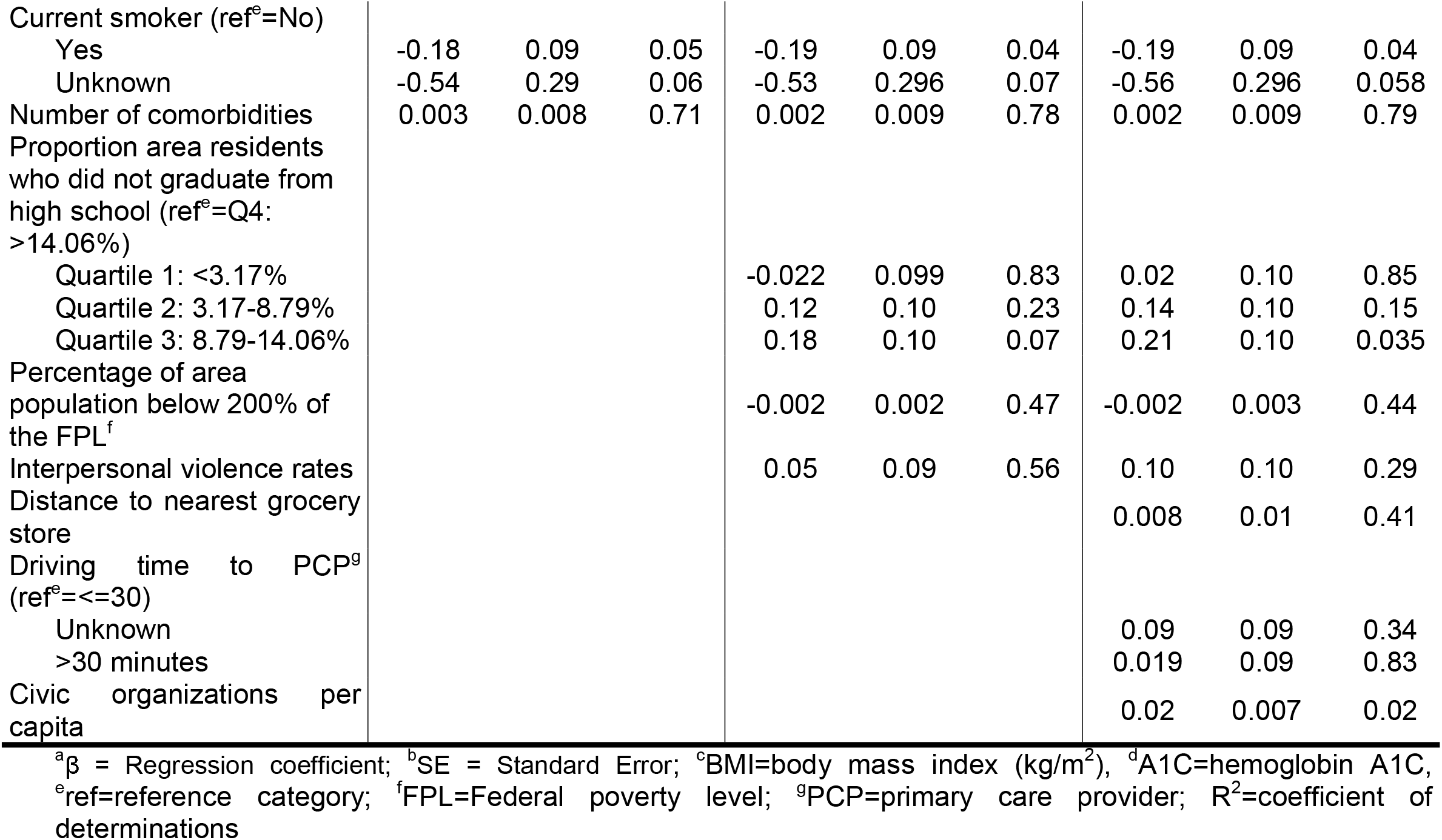
Multivariable models of hemoglobin A1C for 1290 participants with pre-and post-intervention measures.

## Sensitivity Analysis

The results from both the parsimonious model and the mixed-effects model for clinics show no important changes in the parameter estimates for significant associations. See S2-9 Tables.

## Discussion

We hypothesized that individuals from neighborhoods with higher area-level social risks and longer travel time to PCP locations might show less improvement or worsening in cardiometabolic outcomes with clinic-based nurse-led care coordination. Results did not support our hypothesis. We found individuals with higher pre-intervention cardiometabolic measures got worse despite clinic-based nurse-led care coordination. Additionally, women’s LDL-cholesterol worsened compared to men irrespective of pre-intervention levels, and women with high pre-intervention HbA1C (> 6.8%) and BP (>121 mm Hg SBP) got worse compared to men despite clinic-based, nurse-led care coordination. Hence, women with suboptimal or uncontrolled diabetes and hypertension showed worsening of HbA1C and BP, respectively. Patients from areas with higher poverty showed small improvements in BP when adjusted for pre-intervention DBP and in LDL-cholesterol with care coordination. While there were some inconsistent findings across outcomes, the addition of neighborhood-level risks based on geocoded residential addresses did not significantly change associations or fit of models compared to models based on variables that are routinely available in clinical charts.

Gender is the only socially stratifying factor present in >50% of our cohort, and we found female gender was associated with the worsening of all measured cardiometabolic outcomes.[43] Our findings of worsening outcomes in women compared to men are noteworthy because screening and interventions for social determinants of health for adults in primary care have focused little on social determinants largely associated with women, like availability of affordable childcare, adult caregiving, and disparities in employment opportunities for single mothers.[44-49] Moreover, mortality due to cardiovascular diseases has increased in middle-aged women over the last decade.[5, 50, 51] Chronic disease self-management includes 1) Medical management: taking medications, self-monitoring, and lifestyle modifications; 2) Role management: maintaining meaningful life roles with family, friends, and work; 3) Emotional management: coping with anger, fear, frustration, and sadness.[52] Our findings may indicate that individuals, especially women, with suboptimal control of cardiometabolic conditions have additional daily lived behavioral, emotional, and role management contexts, not captured by EHR or neighborhood-level risks, that diminish their ability to benefit from solely clinic-based interventions.[6, 48, 53-56] A study of nationally representative data found that older women had more behavioral and social risks compared to men.[57] Previous studies have shown nurse case management improves diabetes and hypertension control, but increasing baseline measures were associated with worsening of all cardiometabolic measures in our study of nurse-led care coordination.[58, 59] Nurse-led care coordination and chronic disease education may need to be combined with additional tailored disease management components or community-based interventions to improve cardiometabolic outcomes.[58-61]

We found no studies examining patient-level moderating factors of cardiometabolic outcomes with care coordination. An analysis of the Johns Hopkins Community Health Partnership (J-CHiP) program looked at the additional effects of a community-based program addressing social determinants along with nurse case management and found improvement trends in healthcare utilization in Medicaid beneficiaries, but not Medicare beneficiaries.[62]

### Strengths

Our study is the first study to examine the interaction of baseline cardiometabolic risk factors with clinical factors and neighborhood-level social risks to identify individuals who may get less benefit or not benefit from solely clinic-based interventions for CVD prevention. We included variables widely available in clinical charts and neighborhood-level social risks based on patients’ residential addresses to avoid burdening PCPs with additional individual social risk screenings.[24, 25, 63] In addition to variables accounting for most social and behavioral domains recommended by the National Academy of Medicine, our study included 1 factor associated with each of 5 key areas of social determinants of health included in the place-based organizing framework developed by Healthy People 2020, namely, economic stability, education, social and community context, neighborhood environment, and healthcare.[32, 64]

### Limitations of Study

We acknowledge several limitations. First, the cohort consists of Medicaid, Medicare, or dual-eligible beneficiaries of predominantly White, non-Hispanic ethnicity from primary care clinics in central Missouri, which may limit the generalizability of our findings. Second, physical activity is one of the primary CVD risk factors, but we could not identify reliable neighborhood-level measures of physical activity opportunities for our cohort.[65, 66] Third, though burdensome, individual-level social risks, rather than neighborhood-level risks, can improve the ability to predict which patients may benefit from supplementary community-based interventions.[67-69] Lastly, we acknowledge that the LIGHT^2^ care coordination program focused on super-utilizers and reducing care fragmentation rather than reducing health disparities.[27]

## Conclusions

We found increasing cardiometabolic measures and women with uncontrolled baseline cardiometabolic measures were commonly associated with worsening cardiometabolic outcomes in participants of a clinic-based care coordination program. Further research on social and behavioral contexts to understand the causes of these associations is needed. Integration of neighborhood-level risks based on patients’ residential address did not change estimates of associations beyond routinely collected clinical and sociodemographic data in EHR.

## Supporting information

STROBE checklist

## Data Availability

All relevant data are within the manuscript and its Supporting Information files.

## Additional contributions

We are thankful to Ms. Cynthia Haydon for her manuscript editing services.

## Supplemental material

S1 File: Detailed methods for determining area-level social risks and driving time to the grocery store and PCP clinic location. Includes geocoding precision, variables in poverty calculation, variables in educational attainment calculation, area-level variables used in models

S1 Table Multivariable models of Systolic BP for 4618 participants with pre-and post-intervention measures.

S2 Table: Parsimonious model for low-density lipoprotein (LDL) cholesterol

S3 Table: Parsimonious model for HbA1C

S4 Table: Parsimonious model for systolic blood pressure (BP)

S5 Table: Parsimonious model for diastolic BP

S6 Table: Solution for Random Effects by clinics-LDL outcome

S7 Table: Solution for Random Effects by clinics: HbA1C Outcome

S8 Table: Solution for Random Effects by clinics: Systolic BP outcome

S9 Table: Solution for Random Effects by clinics: Diastolic BP outcome

S1 Figure: Interaction of female sex with pre-intervention systolic BP on adjusted change in systolic BP

S2 Figure: Interaction of female sex with pre-intervention systolic BP on adjusted change in diastolic BP

S3 Figure: Interaction of female sex with pre-intervention HbA1C on adjusted change in HbA1C

