## Supplemental Material for "Individual-level and neighborhood-level factors associated with longitudinal changes in cardiometabolic measures in participants of a care coordination program"

S1 File: Detailed methods for determining area-level social risks and driving time to the grocery store and PCP clinic location. Includes geocoding precision, variables in poverty calculation, variables in educational attainment calculation, area-level variables used in models

S1 Table Multivariable models of Systolic BP for 4618 participants with pre-and post-intervention measures.

S2 Table: Parsimonious model for low-density lipoprotein (LDL) cholesterol

S3 Table: Parsimonious model for HbA1C

S4 Table: Parsimonious model for systolic blood pressure (BP)

S5 Table: Parsimonious model for diastolic BP

S6 Table: Solution for Random Effects by clinics- LDL outcome

S7 Table: Solution for Random Effects by clinics: HbA1C Outcome

S8 Table: Solution for Random Effects by clinics: Systolic BP outcome

S9 Table: Solution for Random Effects by clinics: Diastolic BP outcome

S1 Figure: Interaction of female sex with pre-intervention systolic BP on adjusted change in systolic BP

S2 Figure: Interaction of female sex with pre-intervention systolic BP on adjusted change in diastolic BP

S3 Figure: Interaction of female sex with pre-intervention HbA1C on adjusted change in HbA1C

**S1 File:** *Detailed methods for determining area-level social risks and driving time to the grocery store and PCP clinic location*

Data used in the analysis were obtained from available secondary data and spatial analyses of de-identified patient addresses in combination with address-level datasets.

**Geolocating patient records**

To ascertain spatial locations and neighborhood (e.g., census tract) values for analyses, an extract of 9317 LIGHT^2^ patient addresses was created containing only a unique patientID and the patient address (Address, City, County, ZIP Code, and State). All personal health information variables were removed. Records were geocoded or translated from address information to geographic coordinates, providing latitude and longitude attributes. The address file was geocoded using ArcGIS Online World Geocoding Service, version dated June 6, 2019.^1^

Supplemental S1 Table displays the geocoding precision of the resulting dataset. 6507 (69.84%) were located at the housing unit centroid level, (PointAddress, Subaddress), 2691 were located at the street address level (StreetAddress), and 44 were located at the ZIP Code + 4 centroid level (PostalExt). The remaining 0.8% were located at the ZIP code centroid or street name levels. Of 9317 records, over 99% were located with the highest levels of accuracy. Definitions for the geocode precision types can be found by accessing the [ESRI Geocoding Service Output documentation](https://developers.arcgis.com/rest/geocode/api-reference/geocoding-service-output.htm).^1^

| **Geocoding Precision** | | |
| --- | --- | --- |
| **Geocode Precision** | **Count** | **Percentage** |
| PointAddress | 6506 | 69.83 |
| StreetAddress | 2691 | 28.88 |
| PostalExt | 44 | 0.47 |
| StreetName | 43 | 0.46 |
| StreetAddressExt | 20 | 0.21 |
| Postal | 12 | 0.13 |
| Subaddress | 1 | 0.01 |

Census tract and census block group IDs were added to the resulting dataset by performing a spatial intersection between the geocoded results and the 2014 TIGER/Line Block Group Shapefile.^2^ The resulting information was stored in a password protected Microsoft SQL database for further analysis.

**Poverty and education secondary- data elements**

Analysis of variation in patient socio-economic status (SES) required a data source with sub-county variation; this was necessary as 60% of patients were identified as residing in a single county (Boone, Missouri). Data were obtained from the US Census Bureau 2010-2014 American Community Survey (ACS) 5-Year Estimates, accessed through the Census Bureau’s data system.^3^ The ACS is an annual, national survey that contains “social, economic, housing, and demographic characteristics” about the US population. Data are released as 5-year period estimates at units of geography down to the census block group level. From this dataset, 2 variables were selected for inclusion in the analysis: the percentage of census tract residents living below 200% of the federal poverty level (FPL) and the percentage of block group resident adults over age 25 without a high school diploma or equivalency. Percentages were calculated from tabulated data by the University of Missouri Center for Applied Research and Engagement Systems (CARES). Supplemental Table 2 shows the variables used in the poverty calculation:

| **Variables in Poverty Calculation** | | |
| --- | --- | --- |
| **Column ID** | **Table ID & Line Number** | **Definition** |
| 0 | B17024001 | Universe:  Population (for whom poverty status is determined) |
| 1 | B17024003 - B17024010 +  B17024016 - B17024023 +  B17024029 - B17024036 +  B17024042 - B17024049 +  B17024055 - B17024062 +  B17024068 - B17024075 +  B17024081 - B17024088 +  B17024094 - B17024101 +  B17024107 - B17024114 +  B17024120 - B17024127 | Numerator:  All Ages, Ratio of Income to Poverty Level: Under 2.00 |

Supplemental S3 Table shows the variables used in the educational attainment calculation.

| **Variables in Educational Attainment Calculation** | | |
| --- | --- | --- |
| **Column ID** | **Table ID & Line Number** | **Definition** |
| 0 | B15002001 | Universe: Population 25 Years and Over |
| 1 | B15002003-B15002010 + B15002020-B15002027 | Numerator:  Age 25 and older; No schooling completed through 12^th^ grade, no diploma |

The resulting census-tract level poverty and census block group-level educational attainment estimates were added to the de-identified patient dataset by joining the 2 datasets on census tract and block-group IDs.

**Distance data elements**

Variables distance to the nearest grocery store/supermarket and distance to primary care physician clinic location are evaluated at the patient-address level. Distance variables were calculated using an ESRI origin–destination (OD) cost matrix network analysis (ESRI, 2018) using the StreetMap North America (ESRI, 2013) street network dataset.^4^ Patient addresses were assigned as origins, with a search tolerance set to 1200 meters to ensure all records were successfully located along the network dataset.

*Distance to Primary Care Clinic*

Clinic addresses were geocoded using the methods described above and located to the street network dataset. The OD cost matrix network analysis produced a table of 270913 time and distance variables, representing the length and network distance (miles) between each patient and each clinic. This result was filtered to include only those patient + clinic pairs represented in the LIGHT^2^ dataset by cross-referencing a lookup table.

Of 9317 patients in the original geocoded dataset, 841 were not included in the lookup dataset, indicating that there was no assigned primary care clinic address included in the file.

*Distance to Nearest Grocery Store*

Missouri food retailer locations were obtained through the purchase of the ReferenceUSA US Businesses dataset (Infogroup, Inc., 2019).^5^ This database was selected as the research literature shows that the ReferenceUSA (formerly InfoUSA) establishment database has a high degree of accuracy compared to other commercial sources. Small retailers were excluded to approximate stores that sell “a wide variety of healthy foods at affordable prices” per previous studies.^6^ Grocery retailers were defined as establishments with 10 or more employees having a primary SIC code of 541105 (Grocers-Retail) OR primary NAICS code of 445110 (Supermarkets/Other Grocery, Excluding Convenience Strs). This definition excluded certain establishments that sell a wide variety of fruits and vegetables in addition to general merchandise. Therefore, establishments with having a primary NAICS 531102 (Department Stores) with a listed company name of “Target” or “Walmart” were also included in the grocer establishment dataset. The resulting dataset contained 852 geo-located establishments. An OD cost matrix network analysis was again used to determine travel time and distance to each type of food retailer (ESRI, 2018).^4^ The analyses produced matrices of 93167 records (grocery outlets). Results were summarized for each patient to return the minimum values for distance and time, and the cumulative number of destinations within 5-minute binned ranges.

**Physical activity opportunities:**

To assess opportunities for physical activity, we generated WalkScore™ values for each patient’s census block using the WalkScore™ API (WalkScore, 2019). WalkScore™ is a rating (on a scale of 0 to 100) of the pedestrian friendliness of the area around a specified location.^7^ To determine the validity of the WalkScore™ data for our model, we generated a visual map of the WalkScore™ data in Boone county, Missouri. Visual assessment found that addresses in Boone county residential areas with access to safe trails and parks were coded as less/least walkable, indicating that the WalkScore™ dataset may not represent physical activity opportunities in areas with networks of unpaved trails. In addition, WalkScore™ data is missing for most rural locations. Hence, we excluded area-level walkability measures from our models because we could not identify reliable area-level physical activity measures for our suburban and rural patient population cohort.

**Social Capital Measure**

Neighborhood (ZIP-code level) social capital was assessed using the number of civic or social organizations per capita, obtained by summarizing data from the 2017 US Census Bureau ZIP code Business Patterns. Civic or social associations are defined by the North American Industry Classification System (NAICS) economic classification system using the definition established by Rupasingha et al. (2006, with updates).^8^ The social capital and social isolation measures are components of the validated Social Capital Index by Rupasingha et al. (2006, with updates).^8^ Social isolation is measured as percentage of adults living alone at neighborhood-level. However, we included marital status reported in patients’ clinical chart as a variable which indicates 43% of our cohort is married, and we have individual-level single or widowed status for our cohort. Hence we excluded the social isolation measure from our measurement of social capital.

**Domestic Violence Injury Rates:**

Domestic violence injury rates are counts of emergency room (ER) and hospital inpatient discharges for injuries coded as Spouse/partner abuse per 10,000 population. Data were obtained at ZIP code-level from the Missouri Department of Health and Senior Services (DHSS) Missouri Information for Community Assessment (MICA) Injury MICA data profiles query tool. We used multi-year aggregates for years 2011-2015 to avoid data suppression.

| **Area-level variables** | | | |
| --- | --- | --- | --- |
| **Variable Definition** | **Source(s)** | **Source Time Period** | **Geographic Resolution** |
| Percentage of Population below 200% Federal Poverty Level | US Census Bureau, American Community Survey 5-Year Estimates (2014) | 2010-2014 | Census Tract |
| Educational Attainment – Scaled Variable | US Census Bureau, American Community Survey 5-Year Estimates (2014) | 2010-2014 | Census Block Group |
| Travel time to Primary Care > 30 Minutes (Binary) | LIGHT^2^ Patient Dataset  ESRI, StreetMap North America (2013) | 2013-2015  2010 | Address |
| Distance to Nearest Grocery Store | Infogroup, ReferenceUSA (2019) | 2019 | Address |
| Civic and Social Associations Rate (per 100,000) | US Census Bureau, County Business Patterns (2017) | 2017 | ZIP Code |
| Domestic violence injury hospitalization rate (per 1,000) | Missouri Department of Health and Senior Services, Injury MICA (2017) | 2011-2015 | ZIP Code |

| **S1 Table: Multivariable models of systolic blood pressure for 4618 participants with pre-and post-intervention measures** | | | | | | | | | |
| --- | --- | --- | --- | --- | --- | --- | --- | --- | --- |
|  | **Model 1 (R^2^ 0.43)** | | | **Model 2 (R^2^ 0.43)** | | | **Model 3 (R^2^ 0.43)** | | |
| **Parameter** | **Estimate** | **Std Error** | **P-value** | **Estimate** | **Std Error** | **P-value** | **Estimate** | **Std Error** | **P-value** |
| Intercept | 13.47 | 7.82 | 0.08 | -2.096 | 9.62 | 0.83 | -3.17 | 9.66 | 0.74 |
| Pre-intervention BMI^*^ | 0.11 | 0.026 | <.001 | 0.097 | 0.026 | <.001 | 0.09 | 0.026 | <.001 |
| Pre-intervention SBP^†^ | 0.91 | 0.07 | <.001 | 0.93 | 0.07 | <.001 | 0.93 | 0.07 | <.001 |
| Pre-intervention DBP^‡^ | -0.12 | 0.034 | 0.001 | 0.05 | 0.07 | 0.48 | 0.05 | 0.07 | 0.49 |
| Female (ref^\|\|^=male) | -8.16 | 3.95 | 0.04 | -9.06 | 3.95 | 0.02 | -8.93 | 3.95 | 0.02 |
| Pre-intervention SBP^†^ x female sex | 0.061 | 0.03 | 0.04 | 0.067 | 0.029 | 0.02 | 0.067 | 0.03 | 0.02 |
| Non-White race (ref^\|\|^=White) | 0.091 | 0.59 | 0.88 | 0.15 | 0.60 | 0.79 | 0.31 | 0.60 | 0.60 |
| Age | 0.31 | 0.11 | 0.006 | 0.35 | 0.11 | 0.001 | 0.35 | 0.11 | 0.002 |
| Pre-intervention SBP^†^ x age | -0.002 | 0.0009 | 0.02 | -0.002 | 0.0009 | 0.008 | -0.002 | 0.0009 | 0.009 |
| Marital Status (ref^\|\|^=married) |  |  |  |  |  |  |  |  |  |
| Single | -0.48 | 0.57 | 0.40 | -0.51 | 0.58 | 0.38 | -0.42 | 0.58 | 0.47 |
| Widowed | 0.91 | 0.58 | 0.12 | 0.78 | 0.58 | 0.18 | 0.81 | 0.58 | 0.16 |
| Other | 0.72 | 0.62 | 0.24 | 0.50 | 0.62 | 0.42 | 0.54 | 0.62 | 0.39 |
| High healthcare resource utilizer (ref^\|\|^=low utilizer) | -0.19 | 0.57 | 0.74 | -0.14 | 0.57 | 0.80 | -0.11 | 0.57 | 0.85 |
| Mood disorder present (ref^\|\|^=absent or unknown) | -0.26 | 0.51 | 0.60 | -0.29 | 0.51 | 0.57 | -0.26 | 0.51 | 0.61 |
| High risk alcohol use (ref^\|\|^=unknown) | 1.72 | 1.50 | 0.25 | 1.68 | 1.50 | 0.26 | 1.68 | 1.49 | 0.26 |
| Nurse care contacts (ref^\|\|^=Q1: <5) |  |  |  |  |  |  |  |  |  |
| Quartile 2: 5-10 | -0.88 | 0.57 | 0.12 | -0.80 | 0.57 | 0.16 | -0.80 | 0.57 | 0.16 |
| Quartile 3: 11-22 | -0.60 | 0.55 | 0.28 | -0.58 | 0.55 | 0.29 | -0.57 | 0.55 | 0.30 |
| Quartile 4: >22 | -1.18 | 0.56 | 0.03 | -1.19 | 0.56 | 0.03 | -1.17 | 0.56 | 0.04 |
| Current smoker (ref^\|\|^=No) |  |  |  |  |  |  |  |  |  |
| Yes | 0.79 | 0.51 | 0.12 | 0.41 | 0.52 | 0.43 | 0.38 | 0.52 | 0.47 |
| Unknown | -1.30 | 1.77 | 0.46 | -1.44 | 1.77 | 0.41 | -1.49 | 1.77 | 0.40 |
| Number of comorbidities | 0.11 | 0.05 | 0.02 | 0.10 | 0.05 | 0.05 | 0.099 | 0.05 | 0.05 |
| Proportion area residents who did not graduate from high school (ref=Q4: >14.06%) |  |  |  |  |  |  |  |  |  |
| Quartile 1: <3.17% |  |  |  | -1.34 | 0.58 | 0.02 | -1.006 | 0.61 | 0.09 |
| Quartile 2: 3.17-8.79 % |  |  |  | -0.72 | 0.58 | 0.21 | -0.57 | 0.58 | 0.33 |
| Quartile 3: 8.79-14.06% |  |  |  | 0.27 | 0.59 | 0.64 | 0.41 | 0.59 | 0.48 |
| Percentage of area population below 200% of the FPL^#^ |  |  |  | 0.36 | 0.13 | 0.006 | 0.36 | 0.13 | 0.007 |
| Pre-intervention DBP x percentage of population below 200% of the FPL^#^ |  |  |  | -0.005 | 0.002 | 0.007 | -0.005 | 0.002 | 0.008 |
| Domestic violence injury rates |  |  |  | 1.77 | 0.55 | 0.001 | 2.045 | 0.57 | <0.001 |
| Distance to nearest grocery store |  |  |  |  |  |  | 0.043 | 0.06 | 0.47 |
| Driving time to PCP^§^ (ref=<=30) |  |  |  |  |  |  |  |  |  |
| >30 minutes |  |  |  |  |  |  | 0.57 | 0.55 | 0.29 |
| Unknown |  |  |  |  |  |  | 0.53 | 0.54 | 0.33 |
| Civic organizations per capita |  |  |  |  |  |  | 0.06 | 0.04 | 0.17 |
| *BMI=body mass index (kg/m^2^), † SBP=systolic blood pressure, ‡ DBP=diastolic blood pressure, § PCP=primary care provider, \|\| ref=reference category, # FPL=Federal poverty level; **R^2^=coefficient of determinations** | | | | | | | | | |

| **S2 Table: Parsimonious model for LDL cholesterol** (R^2^ = 0.29) | | | |
| --- | --- | --- | --- |
| **Parameter** | **Estimate** | **Standard Error** | **P-value** |
| Intercept | 68.66 | 5.50 | <.001 |
| Pre-intervention BMI^†^ | -0.19 | 0.084 | 0.02 |
| Pre-intervention LDL^*^ | 0.56 | 0.02 | <.001 |
| Female (ref^‡^=male) | 7.76 | 1.30 | <.001 |
| Non-White race (ref=White) | -3.43 | 1.94 | 0.077 |
| Age | -0.26 | 0.05 | <.001 |
| Number of comorbidities | -0.47 | 0.16 | 0.004 |
| Percentage of area population below 200% of the FPL^§^ | -0.14 | 0.045 | 0.002 |
| Interpersonal violence rate | -5.78 | 1.76 | 0.001 |
| Time to PCP^\|\|^<=30 |  |  |  |
| >30 minutes | 0.15 | 1.59 | 0.92 |
| Unknown | -4.27 | 1.91 | 0.025 |
| * LDL=low density lipoprotein, † BMI=body mass index (kg/m^2^), ‡ ref=reference category, § FPL=Federal poverty level, \|\|PCP=primary care provider; **R^2^=coefficient of determinations** | | | |

| **S3 Table: Parsimonious model HbA1C** (R^2^ = 0.39) | | | |
| --- | --- | --- | --- |
| **Parameter** | **Estimate** | **Standard Error** | **P-value** |
| Intercept | 3.73 | 0.41 | <.0001 |
| Pre-intervention BMI^*^ | 0.002 | 0.004 | 0.63 |
| Pre-intervention HbA1C | 0.51 | 0.04 | <.001 |
| Female (ref^†^=male) | -1.29 | 0.34 | <.001 |
| Non-White race (ref^§^=White) | -1.16 | 0.40 | 0.004 |
| Age | -0.006 | 0.003 | 0.05 |
| Current smoker (ref^§^=No) |  |  |  |
| Yes | -0.20 | 0.09 | 0.03 |
| Unknown | -0.58 | 0.29 | 0.05 |
| Pre-intervention A1c x non-White race | 0.14 | 0.06 | 0.01 |
| Pre-intervention A1c x female sex | 0.19 | 0.05 | <.001 |
| Civic organizations per capita | 0.01 | 0.007 | 0.06 |
| Distance to nearest grocery store | 0.01 | 0.009 | 0.25 |
| * BMI=body mass index (kg/m^2^), † A1c=hemoglobin A1c, ‡ PCP=primary care provider, † ref=reference category, \|\| FPL=Federal poverty level; **R^2^=coefficient of determinations** | | | |

| **S4 Table: Parsimonious model for systolic BP** (R^2^ = 0.43) | | | |
| --- | --- | --- | --- |
| **Parameter** | **Estimate** | **Standard Error** | **P-value** |
| Intercept | -3.68 | 8.17 | 0.65 |
| Pre-intervention BMI | 0.096 | 0.03 | <.001 |
| Pre-intervention SBP | 0.95 | 0.07 | <.001 |
| Female (ref=Male) | -7.86 | 3.92 | 0.045 |
| Age | 0.38 | 0.11 | <.001 |
| Pre-intervention SBP x female sex | 0.06 | 0.03 | 0.043 |
| Pre-intervention SBP x age | -0.003 | 0.001 | 0.003 |
| Number of comorbidities | 0.10 | 0.04 | 0.03 |
| Percentage of poverty in the area | 0.30 | 0.06 | <.001 |
| Pre-intervention DBP*Poverty200FPL | -0.004 | 0.0008 | <.001 |
| Interpersonal violence rates | 2.21 | 0.54 | <.001 |
| * BMI=body mass index (kg/m^2^), † SBP=systolic blood pressure, ‡ DBP=diastolic blood pressure, § PCP=primary care provider, \|\| ref=reference category, # FPL=Federal poverty level; **R^2^=coefficient of determinations** | | | |

| **S5 Table: Parsimonious model for Diastolic Blood Pressure (DBP)^a^** (R^2^ = 0.39) | | | |
| --- | --- | --- | --- |
| **Parameter** | **Estimate** | **Standard Error** | **P-value** |
| Intercept | -8.82 | 5.43 | 0.10 |
| Pre-intervention BMI^†^ | 0.078 | 0.01 | <.0001 |
| Pre-intervention SBP^‡^ | 0.15 | 0.04 | <.0001 |
| Pre-intervention DBP^*^ | 0.83 | 0.04 | <.0001 |
| Female (ref^§^=Male) | -5.33 | 2.25 | 0.018 |
| Age | 0.32 | 0.06 | <.0001 |
| Q1: <5 Nurse care contacts (ref^§^=>21) |  |  |  |
| Q2: 5-10 Nurse care contacts | -0.55 | 0.32 | 0.09 |
| Q3: 11-21 Nurse care contacts | -0.07 | 0.31 | 0.81 |
| Q4: >22 Nurse care contacts | -0.87 | 0.32 | 0.006 |
| Smoker Yes (ref=No) | 0.86 | 0.29 | 0.003 |
| Smoking status missing (ref^§^=No) | -1.23 | 1.01 | 0.22 |
| Pre-intervention SBP* Female | 0.04 | 0.017 | 0.01 |
| Pre-intervention SBP *age | -0.003 | 0.0005 | <.001 |
| High risk Alcohol use | 2.43 | 0.85 | 0.004 |
| Number of comorbidities | 0.06 | 0.03 | 0.02 |
| Proportion not graduated HS in the area, Q4 >14.06% (ref^§^) |  |  |  |
| Proportion not graduated HS^\|\|^ in the area, Q1 <3.17% | -0.69 | 0.33 | 0.04 |
| Proportion not graduated HS in the area, Q2 3.17-8.79 % | -0.79 | 0.33 | 0.02 |
| Proportion not graduated HS in the area, Q3 8.79- 14.06% | 0.0002 | 0.33 | 0.99 |
| Percentage of poverty in the area | 0.20 | 0.07 | 0.008 |
| Pre-intervention DBP**Poverty200FPL^#^ | -0.003 | 0.001 | 0.008 |
| Interpersonal violence rates | 0.96 | 0.32 | 0.002 |
| * DBP=diastolic blood pressure, † BMI=body mass index (kg/m^2^), ‡ SBP=systolic blood pressure, § ref=reference category, \|\| HS = High School # FPL=Federal poverty level; **R^2^=coefficient of determinations** | | | |

| **S6 Table: Solution for Random Effects by clinics- LDL outcome** | | | | | | | |
| --- | --- | --- | --- | --- | --- | --- | --- |
| **Effect** | **clinic** | **Estimate** | **Standard Error of Prediction** | | **DF** | **t Value** | **Pr > \|t\|** |
| **clinic** | **Fairview** | -0.17 | 1.20 | | 2035 | -0.14 | 0.88 |
| **clinic** | **Fayette** | 0.088 | 1.45 | | 2035 | 0.06 | 0.95 |
| **clinic** | **Fulton** | -1.89 | 1.25 | | 2035 | -1.5 | 0.13 |
| **clinic** | **Green Meadows** | 0.14 | 1.18 | | 2035 | 0.12 | 0.90 |
| **clinic** | **Keene** | 0.46 | 1.43 | | 2035 | 0.32 | 0.75 |
| **clinic** | **Smiley** | 0.063 | 1.37 | | 2035 | 0.05 | 0.96 |
| **clinic** | **Woodrail** | 1.31 | 1.17 | | 2035 | 1.12 | 0.26 |
| **Covariance Parameter Estimates** | | | | | | | |
| **Covariate Parameter** | | | | **Estimate** | | | |
| **clinic** | | | | 2.48 | | | |
| **Residual** | | | | 916.93 | | | |

| **S7 Table: Solution for Random Effects by clinics: HbA1c Outcome** | | | | | | | |
| --- | --- | --- | --- | --- | --- | --- | --- |
| **Effect** | **clinic** | **Estimate** | **Standard Error of Prediction** | | **DF** | **t Value** | **Pr > \|t\|** |
| **clinic** | **Fairview** | -0.09 | 0.078 | | 1035 | -1.18 | 0.24 |
| **clinic** | **Fayette** | -0.03 | 0.11 | | 1035 | -0.26 | 0.79 |
| **clinic** | **Fulton** | -0.04 | 0.09 | | 1035 | -0.51 | 0.61 |
| **clinic** | **Green Meadows** | 0.17 | 0.07 | | 1035 | 2.3 | 0.02 |
| **clinic** | **Keene** | -0.02 | 0.10 | | 1035 | -0.2 | 0.84 |
| **clinic** | **Smiley** | -0.04 | 0.09 | | 1035 | -0.5 | 0.62 |
| **clinic** | **Woodrail** | 0.06 | 0.08 | | 1035 | 0.73 | 0.46 |
| **Covariance Parameter Estimates** | | | | | | | |
| **Cov Parm** | | | | **Estimate** | | | |
| **clinic** | | | | 0.01469 | | | |
| **Residual** | | | | 1.1994 | | | |

| **S8 Table: Solution for Random Effects by clinics: Systolic BP outcome** | | | | | | | |
| --- | --- | --- | --- | --- | --- | --- | --- |
| **Effect** | **clinic** | **Estimate** | **Standard Error of Prediction** | | **DF** | **t Value** | **Pr > \|t\|** |
| **clinic** | **Fairview** | -2.45 | 0.73 | | 3805 | -3.36 | <.001 |
| **clinic** | **Fayette** | -0.27 | 0.96 | | 3805 | -0.29 | 0.77 |
| **clinic** | **Fulton** | 0.50 | 0.78 | | 3805 | 0.64 | 0.52 |
| **clinic** | **Green Meadows** | -0.07 | 0.68 | | 3805 | -0.1 | 0.92 |
| **clinic** | **Keene** | -0.02 | 0.91 | | 3805 | -0.02 | 0.98 |
| **clinic** | **Smiley** | 1.46 | 0.82 | | 3805 | 1.77 | 0.08 |
| **clinic** | **Woodrail** | 0.86 | 0.68 | | 3805 | 1.26 | 0.20 |
| **Covariance Parameter Estimates** | | | | | | | |
| **Covariate parameter** | | | | **Estimate** | | | |
| **clinic** | | | | 1.9578 | | | |
| **Residual** | | | | 171.92 | | | |

| **S9 Table: Solution for Random Effects by clinics: Diastolic BP outcome** | | | | | | | |
| --- | --- | --- | --- | --- | --- | --- | --- |
| **Effect** | **clinic** | **Estimate** | **Standard Error of Prediction** | | **DF** | **t Value** | **Pr > \|t\|** |
| **clinic** | **Fairview** | -1.91 | 0.46 | | 3806 | -4.13 | <.001 |
| **clinic** | **Fayette** | 0.55 | 0.6038 | | 3806 | 0.92 | 0.36 |
| **clinic** | **Fulton** | 0.15 | 0.4926 | | 3806 | 0.3 | 0.77 |
| **clinic** | **Green Meadows** | 0.33 | 0.4355 | | 3806 | 0.76 | 0.45 |
| **clinic** | **Keene** | 0.12 | 0.57 | | 3806 | 0.21 | 0.83 |
| **clinic** | **Smiley** | 0.44 | 0.5162 | | 3806 | 0.85 | 0.39 |
| **clinic** | **Woodrail** | 0.33 | 0.4382 | | 3806 | 0.74 | 0.46 |
| **Covariance Parameter Estimates** | | | | | | | |
| **Covariate parameter** | | | | **Estimate** | | | |
| **clinic** | | | | 0.883 | | | |
| **Residual** | | | | 57.357 | | | |

**S1 Figure : Interaction of female sex with pre-intervention systolic blood pressure (BP) on adjusted change in systolic BP**

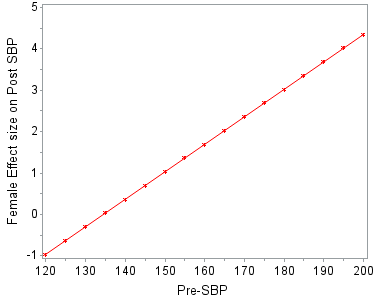

Adjusted change in Systolic BP

Pre-intervention Systolic BP

**S2 Figure:** **Interaction of female sex with pre-intervention systolic blood pressure (BP) on adjusted change in diastolic BP**

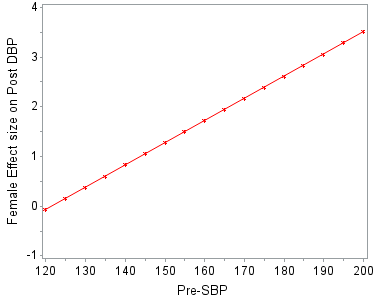

Adjusted change in Diastolic BP

Pre-intervention Systolic BP mm Hg

**S3 Figure: Interaction of female sex with pre-intervention HbA1C on adjusted change in HbA1C**

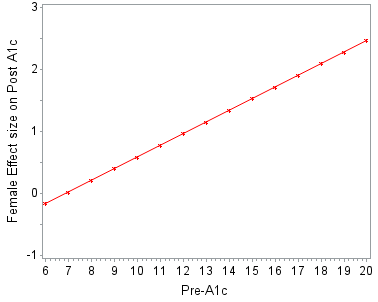

Adjusted change in HbA1C

Pre-intervention HbA1C

4.	ESRI. Arcgis desktop: Release 10. 2018

5.	Infogroup Inc. Referenceusa [dataset]. 2019;2020

6.	Rhone A, Ver Ploeg M, Dicken C, Williams R, Breneman V. Low-income and low-supermarket-access census tracts, 2010-2015. *Economic Information Bulletin No. (EIB-165)*. 2017;2020

7.	Walk Score Professional. Walk score api sign up. 2020;2020

8.	Rupasingha A, Goetz SJ, Freshwater D. The production of social capital in us counties. *The Journal of Socio-Economics*. 2006;35:83-101
