## Supplementary material for "Individual-level and neighborhood-level factors associated with longitudinal changes in cardiometabolic measures in participants of a care coordination program": STROBE checklist

STROBE Statement—Checklist

|  | Item No | Recommendation | Page number |
| --- | --- | --- | --- |
| **Title and abstract** | 1 | (*a*) Indicate the study’s design with a commonly used term in the title or the abstract | Title page and Abstract |
|  |  | (*b*) Provide in the abstract an informative and balanced summary of what was done and what was found | Abstract Page 2 and 3 |
| Introduction | | |  |
| Background/rationale | 2 | Explain the scientific background and rationale for the investigation being reported | Page 4 |
| Objectives | 3 | State specific objectives, including any prespecified hypotheses | Page 4,5 |
| Methods | | |  |
| Study design | 4 | Present key elements of study design early in the paper | Page 5,6 |
| Setting | 5 | Describe the setting, locations, and relevant dates, including periods of recruitment, exposure, follow-up, and data collection | Page 5,6,7 |
| Participants | 6 | (*a*) Give the eligibility criteria, and the sources and methods of selection of participants. Describe methods of follow-up | Page 7,8 |
|  |  | (*b*) For matched studies, give matching criteria and number of exposed and unexposed |  |
| Variables | 7 | Clearly define all outcomes, exposures, predictors, potential confounders, and effect modifiers. Give diagnostic criteria, if applicable | Page 7 to 10, S1 File |
| Data sources/ measurement | 8* | For each variable of interest, give sources of data and details of methods of assessment (measurement). Describe comparability of assessment methods if there is more than one group | Page 8 to 10, S1 File |
| Bias | 9 | Describe any efforts to address potential sources of bias | Page 12 |
| Study size | 10 | Explain how the study size was arrived at | Page 12, Figure 1 |
| Quantitative variables | 11 | Explain how quantitative variables were handled in the analyses. If applicable, describe which groupings were chosen and why | Page 8-12 |
| Statistical methods | 12 | (*a*) Describe all statistical methods, including those used to control for confounding | Page 10-12 |
|  |  | (*b*) Describe any methods used to examine subgroups and interactions | Page 12 |
|  |  | (*c*) Explain how missing data were addressed | Page 7, 8 |
|  |  | (*d*) If applicable, explain how loss to follow-up was addressed |  |
|  |  | (*e*) Describe any sensitivity analyses | Page 12 |
| Results | | |  |
| Participants | 13* | (a) Report numbers of individuals at each stage of study—eg numbers potentially eligible, examined for eligibility, confirmed eligible, included in the study, completing follow-up, and analysed | Figure 1- Flow diagram |
|  |  | (b) Give reasons for non-participation at each stage | Figure 1 |
|  |  | (c) Consider use of a flow diagram | Figure 1 |
| Descriptive data | 14* | (a) Give characteristics of study participants (eg demographic, clinical, social) and information on exposures and potential confounders | Table 1 |
|  |  | (b) Indicate number of participants with missing data for each variable of interest | Figure 1 Flow diagram |
|  |  | (c) Summarise follow-up time (eg, average and total amount) |  |
| Outcome data | 15* | Report numbers of outcome events or summary measures over time | Flow diagram |
| Main results | 16 | (*a*) Give unadjusted estimates and, if applicable, confounder-adjusted estimates and their precision (eg, 95% confidence interval). Make clear which confounders were adjusted for and why they were included | Page 14 to 20, Tables 2,3,4, S1 Table |
|  |  | (*b*) Report category boundaries when continuous variables were categorized | Page 11, 12 |
|  |  | (*c*) If relevant, consider translating estimates of relative risk into absolute risk for a meaningful time period |  |
| Other analyses | 17 | Report other analyses done—eg analyses of subgroups and interactions, and sensitivity analyses | S2-9 tables, S1-3 Figures |
| Discussion | | |  |
| Key results | 18 | Summarise key results with reference to study objectives | Page 21-23 |
| Limitations | 19 | Discuss limitations of the study, taking into account sources of potential bias or imprecision. Discuss both direction and magnitude of any potential bias | Page 23, 24 |
| Interpretation | 20 | Give a cautious overall interpretation of results considering objectives, limitations, multiplicity of analyses, results from similar studies, and other relevant evidence | Page 21 to 24 |
| Generalisability | 21 | Discuss the generalisability (external validity) of the study results | Page 23 |
| Other information | | |  |
| Funding | 22 | Give the source of funding and the role of the funders for the present study and, if applicable, for the original study on which the present article is based | National Center for Advancing Translational Sciences of the National Institutes of Health under Award Number KL2 TR002346. Grant Number 1C1CMS331001-01-00 from the Department of Health and Human Services, Centers for Medicare and Medicaid Services. Publication is solely the responsibility of the authors and do not necessarily represent the official views of the funders |
